# Bayesian approach to comparing the left ventricular volume in myocardial infarction and in normal cases

**DOI:** 10.1101/2021.09.26.21264134

**Authors:** A.T. Hernowo

## Abstract

**Introduction:** Post-contrast delayed-enhancement MRI helps to show the extent of myocardial infarction (MI), as well as allowing the morphometric analysis of the heart structure, e.g., left ventricular volume (LVV). Here the author deployed Bayesian implementation of statistical inference to determine the difference between the LVV in MI cases and in normal controls.

**Methods:** Sixty-seven MI patients and thirty-three controls from the *EMIDEC* dataset challenge were used. These were acquired at the University Hospital of Dijon (France). A cardiologist with 10 years of experience in cardiology and MRI and a biophysicist with 20 years of experience of cardiovascular MRI supervised the acquisition and annotated the images. *ITK-Snap* was used to extract the LVV and Bayesian inference were used to investigate the data.

**Results:** Patients with MI were younger than the controls (58.3 ± 11.5 vs. 66.5 ± 12.9 years; BF_10_ = 17.9). With age taken into consideration, the patients showed larger LVV relative to the controls (128.1 ± 46.3 vs. 83.4 ± 24.4 cm^3^; post-hoc BF_10_ = 12663.8).

**Summary:** Using Bayesian approach, we can conclude decisively that there is volumetric difference or remodeling in individuals with MI.

## I. Introduction

Magnetic resonance imaging (MRI) has been providing non-invasive cardiac imaging and guiding physicians in the management of myocardial infatcion. The widely accepted practice is the application of delayed enhancement post-gadolinium injection T1-weighted imaging [1], with optional T2-weighted acquisition to visualize any edematous tissue. The post-contrast delayed-enhancement MRI (DE-MRI) helps to show the extent of myocardial infarction (MI), as well as allowing the morphometric analysis of the heart structure, e.g., left ventricular volume (LVV).

While group study of either MI or the LVV may have been widely done in various combinations, the approach to the statistical inference are relatively uniform, involving frequentist null-hypothesis significance testing. Nonetheless, Bayesian inference has been getting traction in recent years. [2], [3] There are three main reasons why Bayesian approach is better than the frequentist one: (1) quantifiable evidence in favor of both the null and the alternate hypothesis; (2) prior knowledge utilization; and (3) tangible values of the evidence as more data are accumulated and computed. [4]

Here the author deployed Bayesian implementation of statistical inference to determine the difference between the LVV in MI cases and in normal controls. To show the strength of the evidence supporting either the null or alternate hypothesis, Bayes factor (BF_10_) will be computed.

## II. Methods

Sixty-seven MI patients and thirty-three controls from the automatic Evaluation of Myocardial Infarction from Delayed-Enhancement Cardiac MRI (*EMIDEC)* dataset challenge were used. [5] These were acquired at the University Hospital of Dijon (France). The data were completely anonymized and their acquisition and handling were abiding to the local ethical committee regulation. Since these were compiled retrospectively and completely stripped off their administrative information, it was not necessary to do the process to have ethical approval number.

Siemens MRI scanners (Area (1.5 T) and Skyra (3T)) were used to image the heart during cardiovascular exam with no specific protocol. Short-axis slices of the DE-MRI have been extracted in a retrospective study. The acquisitions taken during breath-hold (ECG-gated), 10 minutes after gadolinium-based contrast agent injection. Resulting MR images were arranged from left ventricular base to apex. Pixel spacing between 1.25 × 1.25 mm^2^and 2 × 2 mm^2^, slice thickness 8 mm, and distance between slices between 8 and 13 mm. The images were provided in NIFTI format. Three screenshot samples ventricular wall and chamber annotation from the controls and the patients group can be seen in the following figure.

A cardiologist with 10 years of experience in cardiology and MRI and a biophysicist with 20 years of experience of cardiovascular MRI supervised the acquisition and annotated the images as seen in figure 1. *ITK-SNAP* was used to extract the LVV and Bayesian inference were used to investigate the data.

**Fig. 1.**
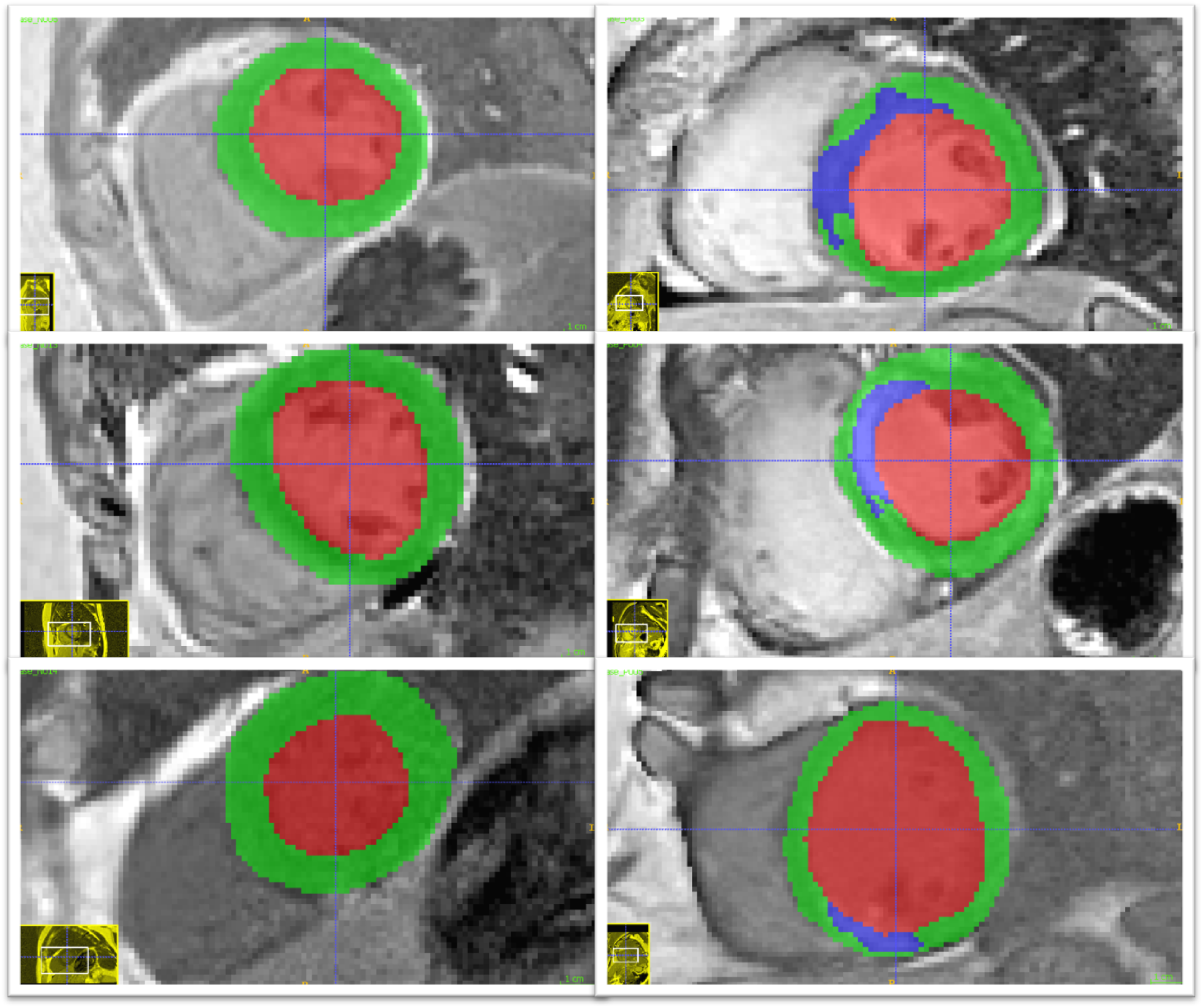
DE-MRI image of the left ventricle. Three random sample left ventricle cross-sectional (mid way between the base and the apex) images are displayed. The controls are on the left side and the patients on the right. The ventricular chamber is annotated with red, while the wall with green and blue colors. Green indicates normally perfused wall, whereas blue indicates the infarction zone.

## III. Results

Mean LVVs of the patients and the controls are described in table 1.

**TABLE I.** Left ventricular volume in controls and patients group.

| Group | N | Mean | SD | SE | 95% CI |  |
| --- | --- | --- | --- | --- | --- | --- |
|  |  |  |  |  | Lower | Upper |
| Controls | 33 | 83.37 | 24.41 | 4.25 | 74.72 | 92.03 |
| Patients | 67 | 128.13 | 46.29 | 5.66 | 116.83 | 139.42 |

Group comparison using Bayesian version of t-test for independent samples revealed that the LVV of the patients is more likely to be different from and larger than the controls (BF_10_=12,663.21; error 2.5·10^−7^%). Effect size (*δ*) parameter estimation is illustrated below.

The larger LVV of the patients is attributed with a large effect size (*δ* = −1.041; 95%*CI* − 1.492 − −0.595-1.041; 95%CI -1.492 - -0.595). To check the robustness of the Bayes factor, varying priors are used and the result can be seen in figure 2.

**Fig. 2.**
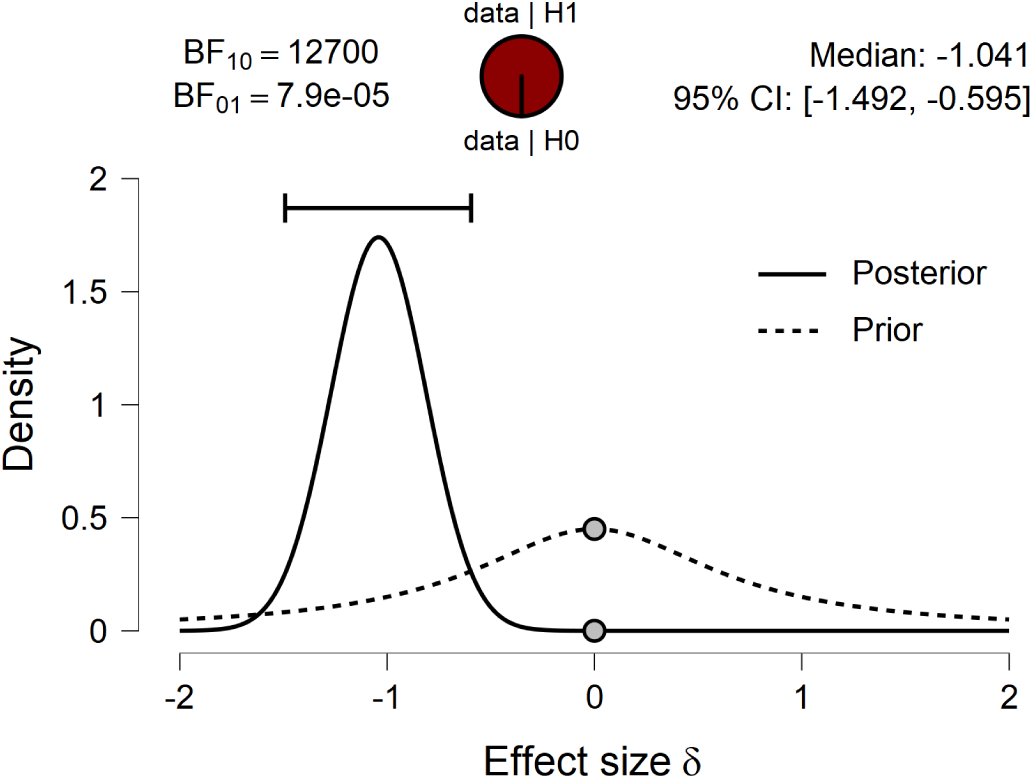
Bayesian two-sample t-test for the parameter *δ*.

The BF_10_ value varies narrowly between 12,663 and 13,507, consistently above 10,000. To see how the Bayes factor vary as more data points were computed, a sequential analysis was done and illustrated in figure 3.

**Fig. 3.**
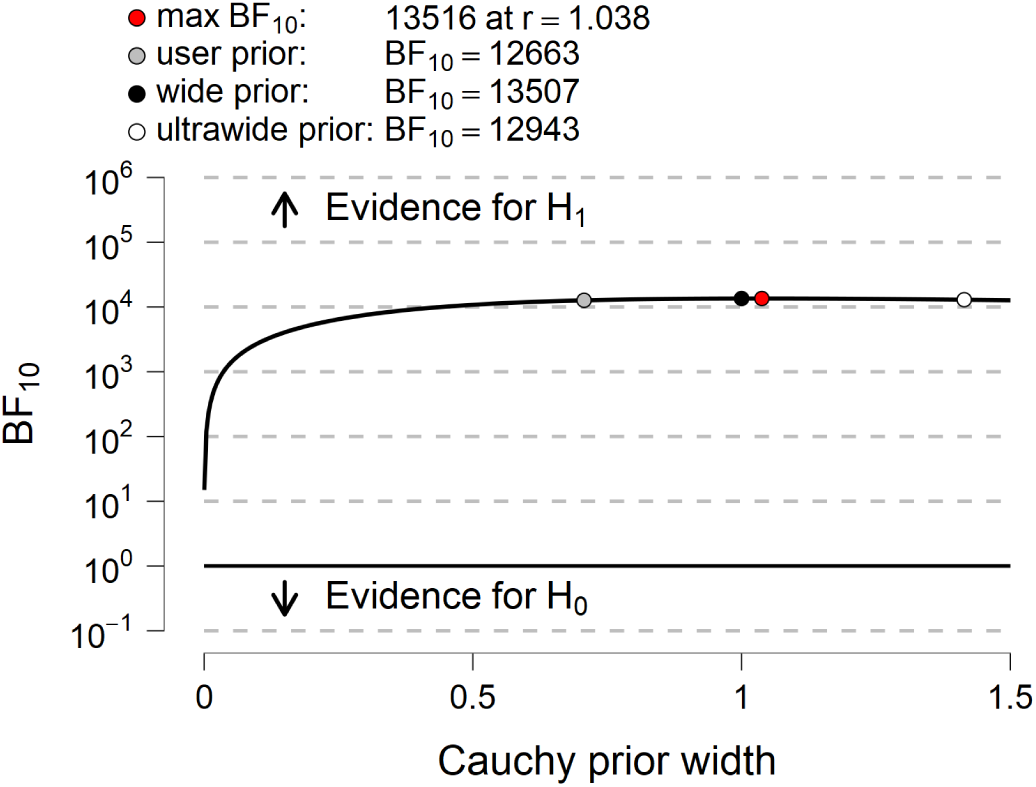
The Bayes factor robustness plot.

The evidential flow for the alternative hypothesis (H_1_) was relatively stable between 10^3^ − 10^4^and hence categorized as an extreme evidence supporting H_1_. To be prudent, we compared the mean age between the two groups. The patients were found to be younger than the controls (58.3 ± 11.5 vs. 66.5 ± 12.9 years; BF_10_ = 17.9; error= 6.777·10^−5^%). Based from the age comparison, it was necessary to take into account when comparing the LVV between the two groups. With age taken into consideration, the patients still showed larger LVV relative to the controls (*post-hoc* BF_10_ = 12,663.84).

## IV. Discussion

The left ventricular volume has been known to increase in patients with myocardial infarction, even as early as 4 weeks after the event. [6], [7], [8], [9] The findings have been confirmed in many studies despite using different modalities to measure the LVV, among which are echocardiography and MRI.

Nonetheless, those reports invariably applied frequentist approach to test their H_0_. It eventually resulted in the acceptance or negation of their H_0_, using *p*-value. The *p*-value itself is proven invaluable to simply nullified the H_0_, but it does not provide any measure of the strength of the evidence backing up or against the alternate hypothesis (H_1_) nor the H_0_. *p*-value is able to show how unlikely the data support the H_0_, but it cannot show how the data support the H_1_. One may, however, argue that the magnitude of difference can be stated as the effect size (*δ*). While *δ* may indicate whether, for example, the mean difference between two groups is small (negligible) or large, *δ* does not indicate the likelihood of the data points to be under or supporting a given hypothesis. Bayes factor on the other hand, indicate a scalar of evidential strength for either the H_0_ or H_1_.

In this report, the author discovered that the LVV in patients is greater than in controls with a BF_10_ of 12,663. Quantitatively, we can say that data are 12,663 times more likely to support the H_1_ rather than the H_0_. In other words, it is extremely likely (>10,000 times more likely) that any LVV measurement in patients with MI will be different (or in this case, greater) from the healthy controls (figure 5).

**Fig. 4.**
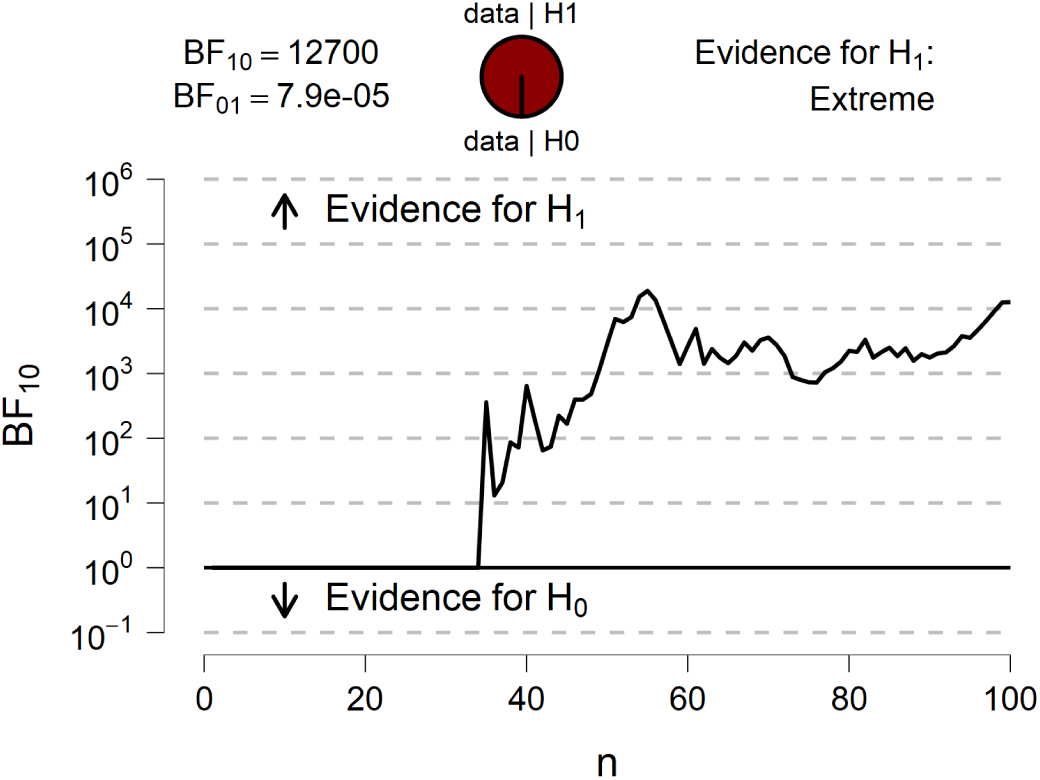
Sequential analysis of the Bayes factor.

**Fig. 5.**
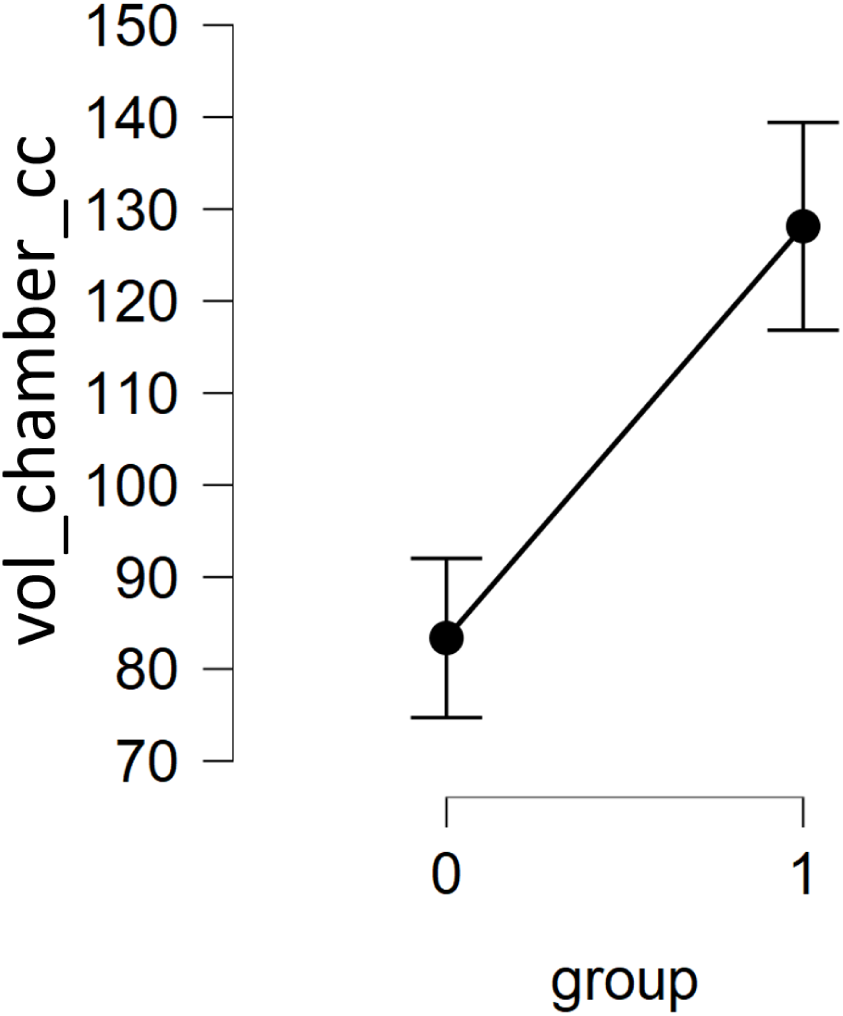
Mean left ventricular chamber volume comparison between controls (“0”) and patients (“1”).

With this kind of results, the author can be cautiously confident that any measurement of the LVV in a patient with MI using this MRI-based volumetry method, will yield a value that will be generally greater than the one of normal individuals of comparable age. Therefore, this finding that the LVV is increased with MI can be considered a general truth. The author recommends more frequent deployment of Bayesian techniques in health or clinical studies to reach quantifiable evidence to build more robust stepping stones for even more further studies.

## Data Availability

The data are from EMIDEC dataset and available online.

